# Suspected Adverse Drug Reactions Associated with Leukotriene Receptor Antagonists Versus First Line Asthma Medications: A National Registry-Pharmacology Approach

**DOI:** 10.1101/2024.06.12.24308833

**Authors:** Mohammed Khan, Christine Hirsch, Alan M. Jones

## Abstract

**Aims:** To determine the suspected adverse drug reaction (ADR) profile of leukotriene receptor antagonists (LTRAs: montelukast and zafirlukast) relative to first-line asthma medications short-acting beta agonists (SABA: salbutamol) and inhaled corticosteroid (ICH: beclomethasone) in the United Kingdom. To determine chemical and pharmacological rationale for the suspected ADR signals.

**Methods:** Properties of the asthma medications (pharmacokinetics and pharmacology) were datamined from the chemical database of bioactive molecules with drug-like properties, European molecular Biology laboratory (ChEMBL). Suspected ADR profiles of the asthma medications was curated from the Medicines and Healthcare products Regulatory Authority (MHRA) Yellow Card interactive drug analysis profiles (iDAP) and concatenated to the standardised prescribing levels (Open Prescribing) between 2018-2023.

**Results:** Total ADRs per 100,000 *R*_*x*_ (*P* < .001) and psychiatric system organ class (SOC) ADRs (*P* < .001) reached statistical significance. Montelukast exhibited the greatest ADR rate at 15.64 per 100,000 *R*_*x*_. The low lipophilic ligand efficiency (LLE = 0.15) of montelukast relative to the controls may explain the promiscuity of interactions with off-target G-coupled protein receptors (GPCRs). This included the dopamine signalling axis, which in combination with bioaccumulation in the cerebrospinal fluid (CSF) to achieve C_max_ beyond a typical dose can be ascribed to the psychiatric side effects observed. Cardiac ADRs did not reach statistical significance but inhibitory interaction of montelukast with the MAP kinase p38 alpha (a cardiac protective pathway) was identified as a potential rationale for montelukast withdrawal cardiac effects.

**Conclusion:** Relative to the controls, montelukast displays a range of suspected system organ class level ADRs. For psychiatric ADR, montelukast is statistically significant (*P* < .001). A mechanistic hypothesis is proposed based on polypharmacological interactions in combination with CSF levels attained. This work further supports the close monitoring of montelukast for neuropsychiatric side effects.

## Introduction

Asthma is a common respiratory condition worldwide. In the UK, 5.4 million people receive treatment, which equates to approximately 1 in 12 adults and 1 in 11 children. [1] This condition is responsible for approximately 3% of primary consultations, 200,000 bed days and 60,000 hospital admissions per year in the UK. [2] Asthma can affect people of all ages, and can range in severity, from very mild, infrequent wheezing, to severe life threatening, closure of the airways. [3] The economic cost of asthma in the UK is > £1bn p.a. which includes medication costs, healthcare professionals’ time, diagnostic tests, and secondary care treatment. [4] The treatment of asthma, involves a stepwise approach, depending on the severity and level interventions required to manage symptoms. [5] Short-acting beta agonists (SABA) for example, salbutamol, is a ‘reliever’ therapy and the first line treatment. If asthma is uncontrolled with a SABA, an inhaled corticosteroid (ICS) for example, beclomethasone is prescribed as maintenance therapy. Should symptoms persist ‘add-ons’ can be given, for example, leukotriene receptor antagonists (LTRA). In the UK, montelukast and zafirlukast are licensed LRTAs (zafirlukast was discontinued in the UK on 31^st^ March 2018). These drugs prevent the pro-inflammatory effects of cysteinyl leukotrienes at their receptors (*CysLT1*) found on the smooth muscles of bronchioles, which helps reduce the levels of inflammation and sensitivity of the airways to asthma triggers. [6]

All drugs can result in adverse drug reactions (ADRs) and can be classified by the DoTS system (dose of the drug, time course of the reaction, and susceptibility factors). [7] **Table S1** highlights the common adverse effects reported with asthma medications prior to this study. It should be noted that ICS side effects in **Table S1** are usually combined with inhalers may cause the same side effects to be reported for ICS so may not be causal for ICS alone. [8] Given the association of neuropsychiatric side effects with the LRTA, montelukast and its circulating metabolites, a pharmaco-epidemiology study is timely to explore the differential ADRs between LRTAs vs SABA vs ICH. [9-14]

Herein, we investigate LRTAs for suspected ADR signals reported between 2018-2023 with salbutamol and beclomethasone as comparators and explore the unique polypharmacology of the drugs for associations with statistically significant suspected ADRs.

## Methods

### Method databases

The Electronic Medicines Compendium (EMC), and the Chemical Database of bioactive molecules with drug-like properties European molecular Biology laboratory (ChEMBL) databases were used to identify the physiochemical properties, pharmacokinetics, and pharmacology data of montelukast, zafirlukast, beclomethasone, salbutamol. [15-18]

### Physiochemical properties

The physicochemical properties included molecular weight, pK_a_, ^*t*^PSA, hydrogen bond donors and acceptors, clogD_7.4_, whether it is a P-glycoprotein substrate and the clog_10_P. The pIC_50_ was calculated using the median IC_50_ of the target *homo sapien* protein for each respective drug. The lipophilic ligand efficacy (LLE) was calculated as follows: LLE = pIC_50_ - clog_10_P (**Table 1**). [18]

**Table 1.** Physiochemical and BBB properties of montelukast, zafirlukast, beclomethasone and salbutamol.

| *Variables* | *Montelukast* | *Zafirlukast* | *Beclomethasone* | *Salbutamol* |
| --- | --- | --- | --- | --- |
| *pIC<sub>50</sub>* | 8.64 | 8.74 | 8.08 | 6.01 |
| *clog<sub>10</sub>P* | 8.49 | 6.4 | 2.15 | 0.34 |
| *LLE* | 0.15 | 2.34 | 5.93 | 5.67 |
| *MW (Da)* | 586.2 | 575.69 | 408.92 | 239.31 |
| *pK<sub>a</sub>* | 4.4 (acidic) | 4.29 (acidic) | 12.44 (neutral) | 9.4 (basic) |
| *tPSA (Å)* | 70.42 | 115.73 | 94.83 | 72.72 |
| *HB acceptors* | 4 | 7 | 5 | 4 |
| *HB donors* | 2 | 2 | 3 | 4 |
| *Clog<sub>10</sub>D<sub>7.4</sub>* | 5.8 | 5.46 | 2.15 | -1.32 |
| *P-glycoprotein Substrate* | Yes | Yes | Yes | Yes |
| *No. of BBB requirements met* | 2 | 1 | 4 | 4 |

### Blood-Brain Barrier (BBB) penetration

The prediction of BBB penetration was by determined by whether the physiochemical properties of the drugs meet requirements and thresholds to allow for penetration. Specifically, molecular weight < 450 Da, ^*t*^PSA <90 Å, neutral or basic characteristics based on *p*Ka, < 6 hydrogen bond donors and < 2 hydrogen bond acceptors, a clogD_7.4_ between 1-3, and ability to be a P-glycoprotein substrate. The more requirements met, the higher predicted BBB penetration. [19]

### Pharmacokinetic properties

The pharmacokinetic properties included bioavailability (*%F*), half-life (*t*_*1/2*_), T_max_ (time to reach the max concentration), C_max_ (peak plasma concentration), whether it was metabolised by a CYP P_450_ enzyme isoform, renal clearance (Cl), volume of distribution (V_d_), total clearance (L/h), and plasma protein binding (PPB). C_max_ was converted from ng/mL to nM. EMC, DrugBank, PubChem, ChEMBL and wider literature were used to find pharmacokinetic data, by searching for “drug name” and “parameter required” (**Table 2**). [15-18, 20-25]

**Table 2.** Pharmacokinetic properties of montelukast, zafirlukast, beclomethasone and salbutamol.

| **Variables** | **Montelukast** | **Zafirlukast** | **Beclomethasone** | **Salbutamol** |
| --- | --- | --- | --- | --- |
| *Bioavailability (%F)* | 64% | 100% | 41% | 24.8% |
| *Half-life (h)* | 2.7 – 5.5 | 8 – 16 | 5.7 | 2.7 – 5.5 |
| *T<sub>max</sub> (h)* | 3 | 3 | 0.7 | 0.17 |
| *C<sub>max</sub> (nM)* | 656.77 | 442.95 | 86.46 | 1678.83 |
| *CYP metabolism* | Yes: CYP3A4, CYP28C, CYP28B | Yes: CYP2C9 | Yes: CYP430 3A | Yes: CYP 450 |
| *Renal excretion* | < 0.2% | 10% | negligible | 272 ± 38 ml/min |
| *Volume of distribution (L)* | 8 - 11 | 70 | 20 | 156 ± 38 (IV) |
| *Clearance (L/h)* | 2.7 | 20 | 150 | 25.22 |
| *PPB (%)* | 99% | 99% | 87% | 10% |

### Pharmacological properties

The ChEMBL database was used to collect the human single protein target bioactivity data for montelukast, zafirlukast, beclomethasone and salbutamol. [18] The median IC_50_ values were used to determine a dependable pIC_50_ value (**Table 3**).

**Table 3.**
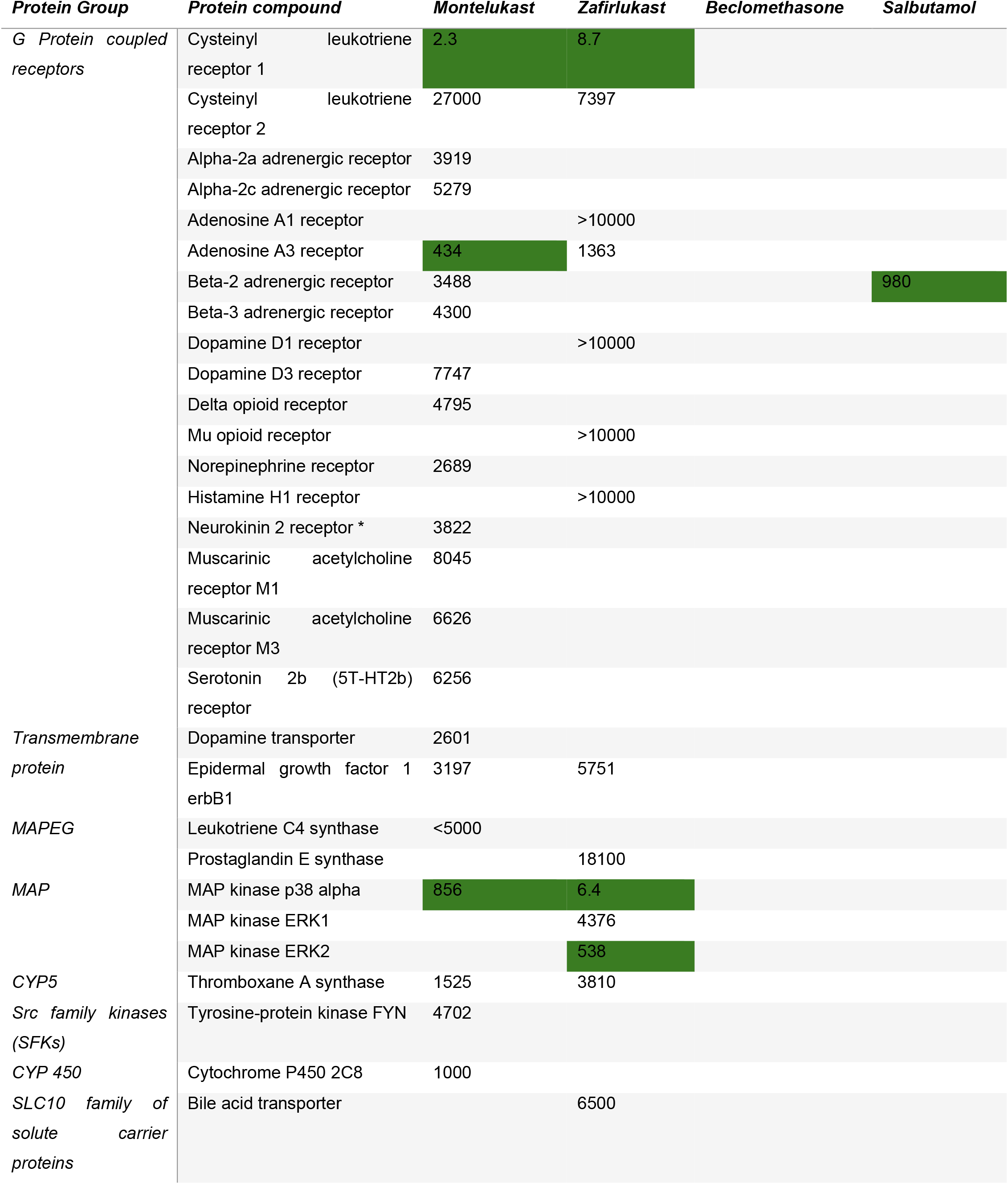
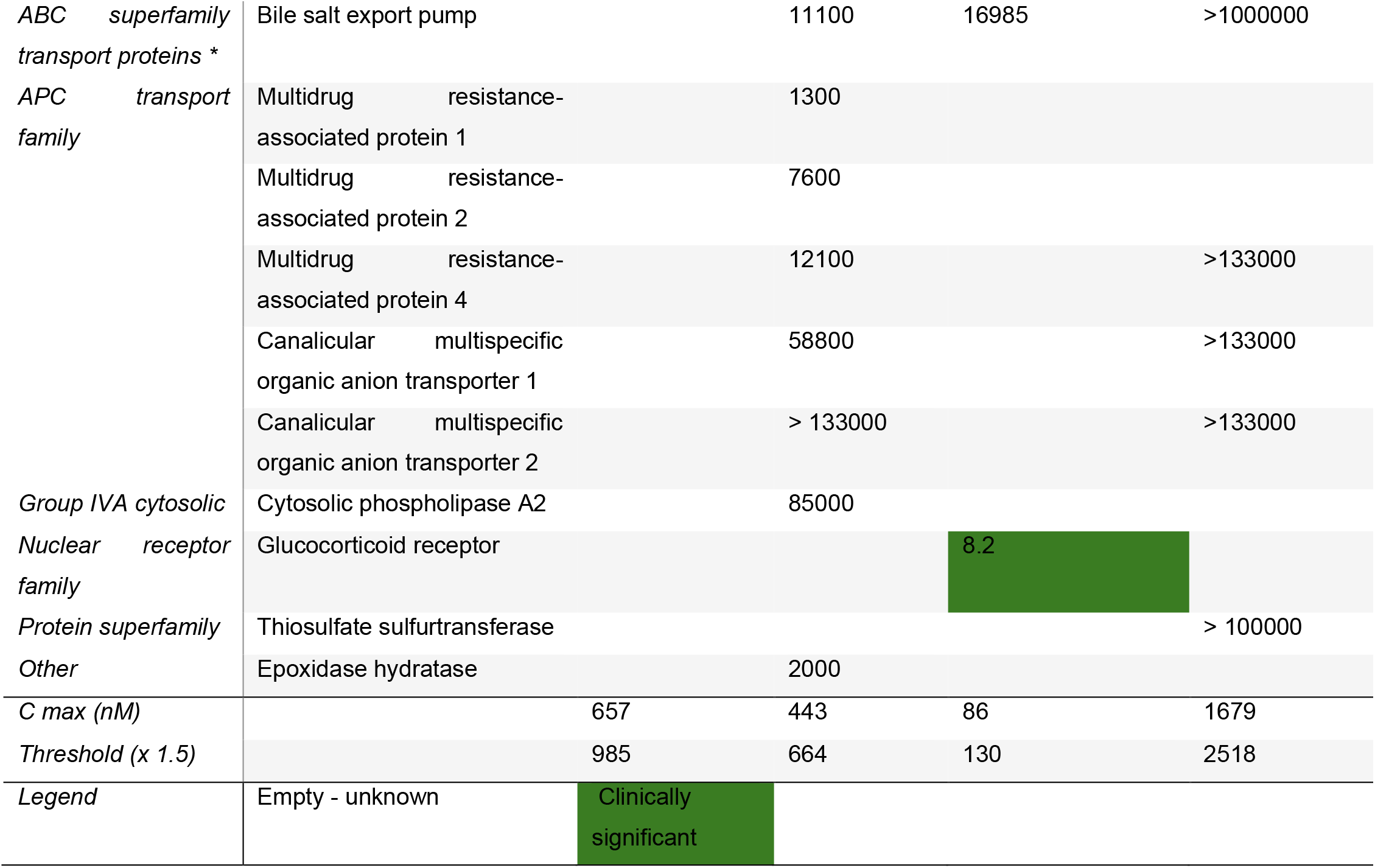
Pharmacology properties of montelukast, zafirlukast, beclomethasone and salbutamol (nM).

### Prescribing data

Prescription data in primary care across England was curated from OpenPrescribing (August 2018 and September 2023). [26] Prescription items (*R*_*x*_) were collated to standardise the suspected adverse drug reactions (ADRs) per 100,000 *R*_*x*_ to factor in variations in prescription rates amongst the medications. A minimum of 100,000 *R*_*x*_ between 2018-2023 was an additional inclusion criterion.

### Adverse drug reactions

Suspected ADR data was datamined from the Medicines and Healthcare products Regulatory Authority (MHRA) Yellow Card interactive Drug Analysis Profile (iDAP). Dates selected were January 2018 to November 2023 for single active constituents only. [27] System organ classes (SOC) with significant ADR rates across the medications and fatalities were selected. The ADR data required standardisation to allow for comparisons between the different drugs. ADRs per 100,000 *R*_*x*_ is a standard approach in signal hypothesis generation [18, 28-33] Drugs were compared to each other in terms of ADR SOCs (**Table S2** and **Table 4**) to determine statistically significant signals and where *P* < .001, high level group terms (HLGT) were additionally explored (**Table S3** and **Table 5**).

**Table 4.** Summary of suspected ADR profiles of montelukast, zafirlukast, beclomethasone and salbutamol in the UK (the number in parentheses represents the standardisation to 100,000 Rx). *P*-values were calculated using a chi-squared test. Supporting data can be found in **Table S2**.

| *Variables* | *Montelukast* | *Zafirlukast* | *Beclomethasone* | *Salbutamol* | *P* |
| --- | --- | --- | --- | --- | --- |
| **Total prescriptions** | 18757214 | 1327 | 70250968 | 107619145 | - |
| **Total ADRs** | 2935 (15.64) | 9 (678.22) | 2327 (3.37) | 1342 (1.24) | < .001 |
| **Fatalities** | 2 (0.01) | 0 (0) | 1 (0) | 15 (0.01) | .95 |
| **Cardiac disorder** | 26 (0.13) | 0 (0) | 71 (0.1) | 68 (0.06) | .98 |
| **Fatalities** | 0 (0) | 0 (0) | 0 (0) | 0 (0) | - |
| *Palpitations* | 23 (0.12) | 0 (0) | 59 (0.08) | 24 (0.02) | .96 |
| **Gastrointestinal disorder** | 204 (1.08) | 0 (0) | 221 (0.31) | 75 (0.06) | .56 |
| **Fatalities** | 0 (0) | 0 (0) | 0 (0) | 0 (0) | - |
| *Abdominal pain* | 17 (0.09) | 0 (0) | 3 (0.00) | 1 (0) | .92 |
| *Nausea* | 39 (0.20) | 0 (0) | 37 (0.05) | 9 (0) | .88 |
| **Nervous system disorder** | 322 ((1.71) | 1 (72.88) | 303 (0.43) | 154 (0.14) | .39 |
| **Fatalities** | 0 (0) | 0 (0) | 0 (0) | 0 (0) | - |
| *Headache* | 72 (0.38) | 0 (0) | 67 (0.09) | 19 (0.01) | .79 |
| **Psychiatric disorder** | 1542 (8.22) | 0 (0) | 174 (0.24) | 73 (0.06) | < .001 |
| **Fatalities** | 2 (0.01) | 0 (0) | 0 (0) | 1 (0) | .99 |
| *Agitation* | 34 (0.18) | 0 (0) | 6 (0) | 1 (0) | .84 |
| *Anxiety* | 165 (0.87) | 0 (0) | 35 (0.04) | 7 (0) | .46 |
| *Depression* | 82 (0.43) | 0 (0) | 7 (0) | 4 (0) | .66 |
| *Hallucinations* | 42 (0.22) | 0 (0) | 1 (0) | 3 (0) | .80 |
| *Nightmare* | 162 (0.86) | 0 (0) | 1 (0) | 0 (0) | .42 |
| *Suicidal ideation* | 62 (0.33) | 0 (0) | 2 (0) | 2 (0) | .72 |
| *Completed suicide* | 3 (0.01) | 0 (0) | 0 (0) | 0 (0) | - |
| ***Respiratory, thoracic, and mediastinal disorders*** | 125 (0.66) | 0 (0) | 587 (0.83) | 528 (0.24) | .85 |
| ***Fatalities*** | 0 (0) | 0 (0) | 0 (0) | 0 (0) | - |

**Table 5.** Psychiatric System Organ Class> High Level Group Term (HLGT). Order of information presented: raw number of suspected reports, standardised to prescribing level in parentheses, fatalities in square brackets. HLGT reported where at least one drug has n > 1 report. *P* values reported where there were *n* > 3 reports. Supporting data can be found in **Table S3**.

| ***HLGT*** | **Montelukast** | **Beclomethasone** | **Salbutamol** | ***P*** |
| --- | --- | --- | --- | --- |
| Anxiety disorders and symptoms | 275 (1.47) | 58 (0.08) | 22 (0.02) | 0.28 |
| Changes in physical activity | 37 (0.20) | 8 (0.01) | 1 (0.00) | - |
| Cognitive and attention disorders and disturbances | 6 (0.03) | 1 (0.00) | 0 | - |
| Communication disorders and disturbances | 15 (0.08) | 1 (0.00) | 0 | - |
| Deliria (incl confusion) | 19 (0.10) | 5 (0.01) | 3 (0.00) | 0.92 |
| Depressed mood disorders and disturbances | 162 (0.86) | 17 (0.02) | 8 (0.01) | 0.45 |
| Developmental disorders NEC | 1 (0.01) | 0 | 0 | - |
| Dissociative disorders | 5 (0.03) | 0 | 0 | - |
| Disturbances in thinking and perception | 76 (0.41) | 5 (0.01) | 6 (0.01) | 0.69 |
| Impulse control disorders NEC | 4 (0.02) | 0 | 0 | - |
| Manic and bipolar mood disorders and disturbances | 6 (0.03) | 0 | 0 | - |
| Mood disorders and disturbances NEC | 182 (0.97) | 25 (0.04) | 5 (0.00) | 0.41 |
| Personality disorders and disturbances in behaviour | 144 (0.77) | 14 (0.02) | 1 (0.00) | - |
| Psychiatric and behavioural symptoms NEC | 65 (0.35) | 7 (0.01) | 1 (0.00) | - |
| Psychiatric disorders NEC | 26 (0.14) | 0 | 6 (0.01) | - |
| Schizophrenia and other psychotic disorders | 12 (0.06) | 0 | 1 (0.00) | - |
| Sexual dysfunctions, disturbances, and gender identity disorders | 0 | 0 | 1 (0.00) | - |
| Sleep disorders and disturbances | 411 (2.19) | 30 (0.04) | 15 (0.01) | 0.12 |
| Somatic symptom and related disorders | 0 | 0 | 1 (0.00) | - |
| Suicidal and self-injurious behaviours NEC | 96 (0.51) [2] | 3 (0.00) | 2 (0.00) | 0.60 |

## Results

### Molecular properties

The structures of the medications studied are shown in **Figure 1**.

**Figure 1.**
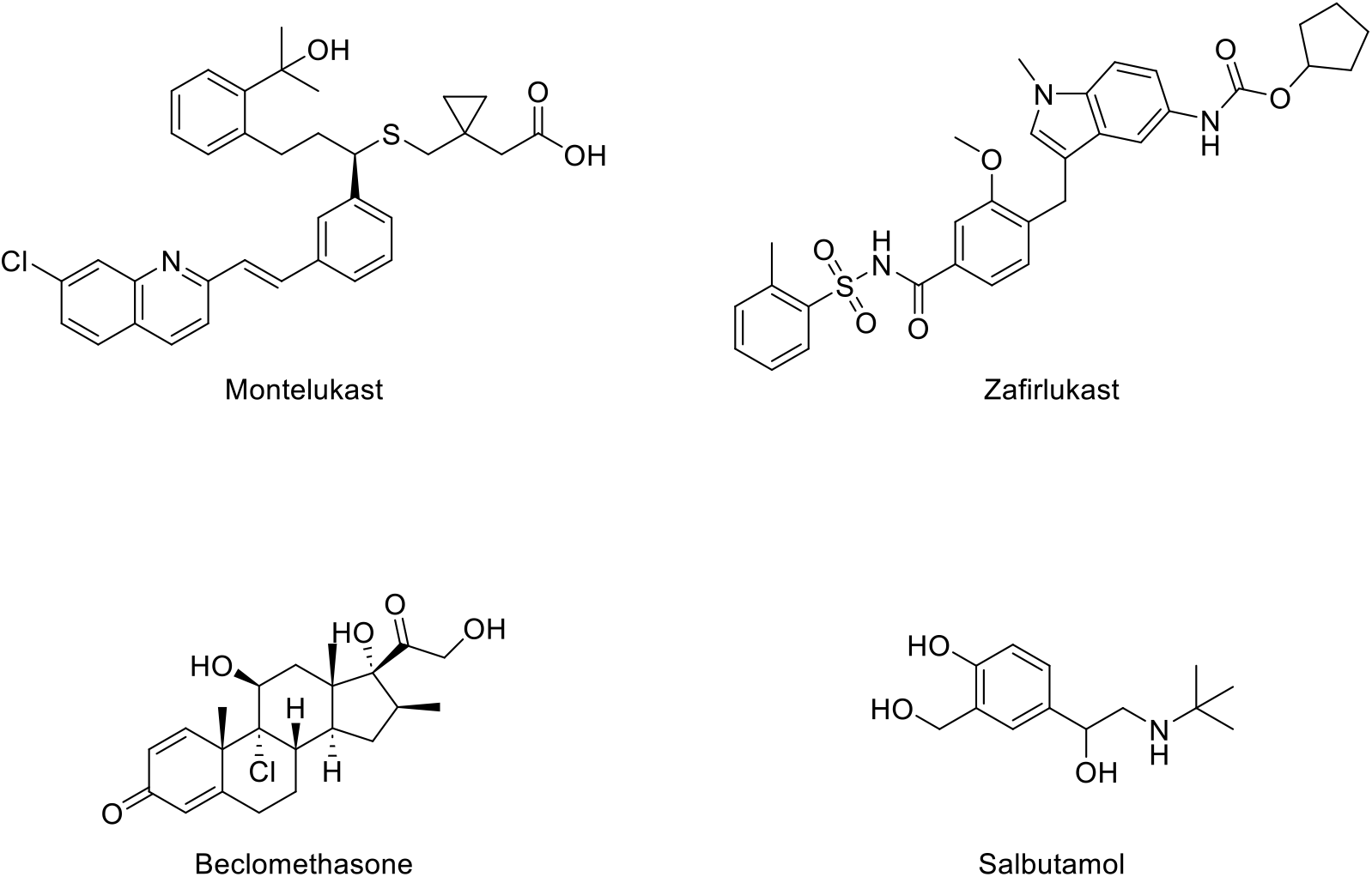
Structures of asthma medications studied (LRTAs: montelukast and zafirlukast, controls ICS: beclomethasone and SABA: salbutamol).

The physiochemical properties of montelukast, zafirlukast and the controls beclomethasone and salbutamol are displayed in **Table 1**.

Montelukast was the most lipophilic drug (clog_10_P = 8.49) and zafirlukast is the most potent (pIC_50_ = 8.74). Montelukast and zafirlukast both had an LLE significantly below 5 (0.15 and 2.34 respectively). The properties which met the requirements for BBB penetration are highlighted in green (**Table 1**). Salbutamol and beclomethasone met both the molecular weight requirement (<450 Da) and are either neutral or basic. Montelukast and salbutamol had a ^*t*^PSA < 90 Å (70.42 Å and 72.72 Å respectively). No drugs had less than 2 hydrogen bond acceptors, however, all drugs had less than 6 hydrogen bond donors. Only beclomethasone had a clogD_7.4_ between 1 and 3 (2.15). All drugs are P-glycoprotein substrates, but this mechanism declines with age.

### Pharmacokinetic properties

The pharmacokinetic properties of the LTRAs and the controls are shown **Table 2**. Zafirlukast had the highest bioavailability of *F =* 100% (oral), while the lowest was salbutamol at 24.8%. The T_max_ was the same for montelukast and zafirlukast (3 h) but lower for beclomethasone and salbutamol (0.7 h and 0.17 h, respectively). All drugs were metabolised by cytochrome P_450_ (CYP) familial isoforms. The V_d_ of salbutamol was highest (143.2 L) and the lowest was montelukast (8-11 L). Montelukast and zafirlukast had high plasma protein binding (PPB) of 99% compared to that of salbutamol at 10%.

### Pharmacology properties

The pharmacology results show the clinically significant values in green with an inhibitory value below the C_max_ threshold (**Table 3**). A threshold was created (x1.5) to include off-target pharmacological activities that may fall outside the C_max_ due to differential dosing regimens. Montelukast was the most potent towards its target receptor, cysteinyl leukotriene receptor 1 (2.3 nM), then zafirlukast (8.7 nM). Montelukast showed potent interaction with off-target, adenosine A3 receptor (43 nM). Both montelukast and zafirlukast were potent towards off-target, MAP kinase p38 alpha (856 nM and 6.5 nM, respectively). Zafirlukast interacted with off-target, MAP kinase ERK2 (538 nM). Salbutamol was potent towards its on-target protein beta-2 adrenergic receptor (980 nM). Beclomethasone was potent towards its on-target receptor, glucocorticoid receptor (8.2 nM). Montelukast and zafirlukast were found to have potent interactions with additional off-target proteins (**Table 3**).

### ADRs

Total ADRs and psychiatric ADRs were found to be statistically significant at SOC level (**Table 4**). Further exploration of psychiatric ADRs at HLGT level are shown in **Table 5**. Montelukast provided a much higher ADR rate per 100,000 *R*_*x*_ in multiple organ class categories including cardiac disorder 0.13 (compared to 0.1 and 0.06 beclomethasone and salbutamol respectively), gastrointestinal disorder (1.08), nervous system disorders (1.71) and psychiatric disorders (8.22) per 100,000 *R*_*x*_. Montelukast was the only drug with a completed suicide ADR rate at 0.01 per 100,000 *Rx*, whereas the others were zero. **Table 4** also shows that montelukast had a higher respiratory, thoracic, and mediastinal disorder than both beclomethasone and salbutamol but did not reach significance.

## Discussion

### Total ADRs and fatalities

Zafirlukast was discontinued in 2018 due to commercial reasons. [34] This led to the drug not meeting the requirements for the inclusion criteria for the ADR/prescribing data collection as it had less than 100,000 *Rx* and therefore excluded from the discussion of addressing ADRs and fatalities.

Amongst all the drugs, montelukast had the highest ADR rate at 15.64 per 100,000 *R*_*x*_, followed by beclomethasone at 3.37 and salbutamol at 1.2. Montelukast had significantly lower LLE compared to both beclomethasone and salbutamol (0.15 compared to 5.93 and 5.67) (**Table 1**). This implies greater promiscuity, and the data corroborates this, as montelukast potently inhibits two off-target proteins (**Table 3**). The highest rate for fatalities across the drugs was shared by both montelukast and salbutamol (0.01 per 100,000 *R*_*x*_).

### Psychiatric ADRs

All SOC ADRs analysed were found to not reach statistically significant except for psychiatric ADRs (**Table 4**). Montelukast (*P* < .001 provided the highest ADR rate at 8.22 per 100,000 *R*_*x*_, which is logarithmically higher than the controls, beclomethasone, and salbutamol (0.24 and 0.06 per 100,000 *R*_*x*_, respectively).

Further data-mining beyond SOCs to HLGTs (**Table 5**) revealed emerging trends between montelukast and the controls but in all cases did not reach statistical significance (*P* > .001). Anxiety disorders and symptoms (275 vs 70 reports for montelukast vs controls respectively), depressed mood disorders (162 vs 25 reports for montelukast vs controls respectively), disturbances in thinking and perception (76 vs 11 reports for montelukast vs controls respectively), mood disorders (182 vs 30 reports for montelukast vs controls respectively),, sleep disorders (411 vs 45 reports for montelukast vs controls respectively), and suicidal and self-injurious behaviours (96 vs 5 reports for montelukast vs controls respectively), showed clear trends with higher reports for montelukast for both directly suspected ADRs and when standardised to prescribing levels compared to the controls. Furthermore, two fatalities within the suicidal and self-injurious category emerged for montelukast compared to the controls (0 for both salbutamol and beclomethasone).

A previous study compared the side effects of antidepressants and immune modulators including montelukast, in human cell lines. Montelukast had a similar side effect profile to the atypical antidepressant, bupropion, which is a dopamine and norepinephrine reuptake inhibitor. [35] Montelukast impacts dopamine levels at a cellular level, despite not being pharmacologically relevant at the protein level based on a single dose C_max_ (**Table 3**). However, physiochemical data can only predict to a certain extent. Another study found that montelukast (at the recommended dose of 10 mg) was discovered in the cerebrospinal fluid (CSF) at a therapeutic dose. The central nervous system (CNS) was then evaluated afterwards to show that montelukast had penetrated the BBB significantly which differs from the predicted BBB in **Table 1**. [36] Furthermore a study of neuropsychiatric events in children, adolescents, and young adults taking montelukast found that montelukast accumulates in the CSF. [37]. This accumulation at levels sufficient to impact the dopamine receptor (7.7 µM, **Table 3**) may potentially explain some of the psychiatric side effects.

### Cardiac ADRs and fatalities

Montelukast had a higher cardiac ADR rate at 0.13 per 100,000 *R*_*X*_ compared to both beclomethasone and salbutamol (**Table 4**). Suspected montelukast induced heart palpitations, were significantly higher (0.12) in comparison with beclomethasone and salbutamol (0.08 and 0.02 per 100,000 *R*_*x*_).

Montelukast is only usually added to asthma treatment if a patient has not responded to first line treatment with salbutamol with regular low dose inhaled corticosteroid (beclomethasone) has not been successful as an alternative to an increased dose of inhaled corticosteroid, allowing management of symptoms via an alternative pharmacological pathway. [38-39]

Montelukast has off target interactions with Adenosine A3 receptor and MAP kinase p38 alpha. The interaction with MAP kinase p38 alpha may be a cause of the cardiac effects. The p38 MAPK pathway plays an integral role in cardiac contractility and cardiac remodelling, and thus inhibiting this protein may be a potential cause for the cardiac ADRs resulting with montelukast. [40] Cessation of montelukast was considered a possible trigger in a 53-year-old male endurance athlete who presented with a two-week history of symptomatic ventricular ectopics, confirmed by ECG and 24-hour tape. Other investigations were considered normal with no alternative identifiable causes for the arrhythmias which resolved 24 hours after re-starting the montelukast. [41]

Montelukast also inhibits adenosine A3 receptor this has a significant role in myocardial infarction and atherosclerosis. Adenosine A3 receptor has cardioprotective characteristics which montelukast inhibits, which may make patients susceptible to deleterious cardiac affects. [42]

### Gastrointestinal disorders and site of administration ADRs

Montelukast exceeded both beclomethasone and salbutamol’s suspected gastrointestinal (GI) ADR rates per 100,000 *R*_*x*_ at 1.08 versus 0.31 and 0.06, respectively (**Table 4**). Montelukast caused a greater portion of GI ADRs which may be attributed to montelukast containing a lactose formulation, which certain patients may not respond well to.

There are directions for prescribers to assess whether the patient can tolerate this montelukast formulation by considering hereditary lactose conditions. [15] No fatalities were observed during this study for GI ADRs with non-serious ADRs reported. For abdominal pain, montelukast had a rate of 0.09 per 100,000 *R*_*x*_ whereas beclomethasone and salbutamol had zero. This is consistent to known side effects with montelukast (**Table S1**), therefore corroborating the validity of the approach using Yellow Card registry data. [8]

### Nervous system ADRs

Montelukast had the highest ADR rate at 1.71 per 100,000 *R*_*x*_, whereas beclomethasone and salbutamol were lower (0.43 and 0.14, respectively). These ADRs share a similar background to psychiatric ADRs with BBB penetrability involved and affecting the dopamine axis. [35-36] Headaches are associated with montelukast at 0.38 per 100,000 Rx whereas for 0.09 and 0.01 for salbutamol and beclomethasone which was validated with comparison to known side effects of montelukast. [8]

## Limitations

A limitation of this study is that zafirlukast was discontinued in 2018 resulting in only 1 year of prescribing data during the 5-year timeline of this study. The reason for the discontinuation of zafirlukast was reported as distribution issues rather than safety concerns of the drug.

The MHRA Yellow Card interactive Drug Analysis Profile provides all the suspected ADRs that have been self-reported by both healthcare professionals and patients which is known to lead to a high degree of underreporting of side effects. This underreporting reduces the power of comparisons made across the drugs.

It should also be noted that some drugs are in the public mindset (e.g. montelukast) when announcements are made regarding potential side effects, this may cause an uptick in reports generated, in comparison to a drug (e.g. salbutamol) which is a well-established first line treatment that may not be reported at the same rate.

Causality does not need to be established before reporting to the Yellow Card scheme, therefore, the drug in question may not be the cause of the effect but rather other factors (polypharmacy, QoL, comorbidity), leading to the misrepresentation of the ADRs. Consequently, an assertive claim about the drug and which ADR it may cause, based alone on the MHRA yellow card scheme is not possible. However, trends and signal detection are possible.

ADRs used in this study are reported between January 2018 and November 2023, whilst the prescription data was available from August 2018 to September 2023. This does not affect the standardisation of drugs as the time frame remains consistent but may not appropriately reflect the drugs across the full 5 years of the study.

## Conclusion

Based on the data observed in this study, montelukast has shown a wide range of suspected ADRs, ranging from cardiac, gastrointestinal, nervous system, and psychiatric effects. The physiochemical data showed a poor prediction of the BBB penetration of montelukast, but evidence from the literature suggest that montelukast does penetrate the BBB and accumulate in the CSF.

The research approach employed in this study was validated with the rediscovery of known montelukast ADRs. In particular, the elevated levels of nervous system and psychiatric disorders reported with montelukast are consistent with the MHRA field notice about adverse effects associated with montelukast. [43]. Montelukast has been associated with mood changes, depression, aggression, hallucinations, and suicidal thoughts and this work further supports these neuropsychiatric findings and a potential pharmacological rationale via interaction with the dopamine GPCR axis through CSF accumulation.

Cardiac ADRs associated with montelukast have a tentative relationship with the pharmacological profile which supports the inhibition of off-target proteins that have an association cardiac ADRs.

The research demonstrated a new perspective that combines pharmacovigilance registries and chemical-pharmacological data to understand the potential causes of the ADRs of LTRAs.

## Data Availability

All data produced in the present work are contained in the manuscript and supporting materials.

https://yellowcard.mhra.gov.uk/idaps

## Acknowledgements

The authors thank MHRA, OpenPrescribing, and ChEMBL for the publicly accessible curated open-access data that was used in this study.

