## Supplementary Information for "Suspected Adverse Drug Reactions Associated with Leukotriene Receptor Antagonists Versus First Line Asthma Medications: A National Registry-Pharmacology Approach"

### CONTENTS

| | |
|---|---|
| **Table S1** | **Page 2** |
| **Table S2** | **Page 3-4** |
| **Table S3** | **Page 5-12** |

**Table S1.** Known side effects of asthma medication.

| Short-acting beta-2 agonist | Inhaled Corticosteroid | Leukotriene receptor antagonist |
|---|---|---|
| Arrythmias<br>Fine tremor<br>Headache<br>Tachycardia | Arrythmias<br>Fine tremor<br>Headache<br>Tachycardia | Aggressive behaviour<br>Anxiety<br>Headache<br>Hallucinations<br>Sleeping issues<br>Abdominal pain |

**Table S2.** Raw system organ class ADRs/fatalities and relative to 100,000 $R_x$ (indicated by (s).

| | Montelukast | | | | Zafirlukast | | | | Beclomethasone | | | | Salbutamol | | | |
|---|---|---|---|---|---|---|---|---|---|---|---|---|---|---|---|---|
| SOC | All | All (s) | Fatal | Fatal (s) | All | All (s) | Fatal | Fatal (s) | All | All (s) | Fatal | Fatal (s) | All | All (s) | Fatal | Fatal (s) |
| Total | 2935 | 15.6473131 | 2 | 0.010663 | 9 | 678.2216 | 0 | 0 | 2372 | 3.376466 | 1 | 0.001423 | 1342 | 1.24699 | 15 | 0.013938 |
| Blood and lymphatic blood disorder | 9 | 0.04798154 | 0 | 0 | 0 | 0 | 0 | 0 | 5 | 0.007117 | 0 | 0 | 2 | 0.001858 | 0 | 0 |
| Cardiac disorders | 26 | 0.13861334 | 0 | 0 | 0 | 0 | 0 | 0 | 71 | 0.101066 | 0 | 0 | 68 | 0.063186 | 0 | 0 |
| Congenital, familial and genetic disorders | 3 | 0.01599385 | 0 | 0 | 0 | 0 | 0 | 0 | 1 | 0.001423 | 0 | 0 | 4 | 0.003717 | 0 | 0 |
| Ear and labyrinth disorders | 9 | 0.04798154 | 0 | 0 | 0 | 0 | 0 | 0 | 11 | 0.015658 | 0 | 0 | 5 | 0.004646 | 0 | 0 |
| Endocrine disorders | 3 | 0.01599385 | 0 | 0 | 0 | 0 | 0 | 0 | 3 | 0.00427 | 0 | 0 | 1 | 0.000929 | 0 | 0 |
| Eye disorders | 22 | 0.11728821 | 0 | 0 | 0 | 0 | 0 | 0 | 85 | 0.120995 | 0 | 0 | 24 | 0.022301 | 0 | 0 |
| Gastrointestinal disorders | 204 | 1.08758156 | 0 | 0 | 0 | 0 | 0 | 0 | 221 | 0.314586 | 0 | 0 | 75 | 0.06969 | 0 | 0 |
| General disorders and administration site conditions | 242 | 1.29017028 | 0 | 0 | 2 | 150.7159 | 0 | 0 | 251 | 0.35729 | 1 | 0.001423 | 206 | 0.191416 | 9 | 0.008363 |
| Hepatobiliary disorders | 4 | 0.02132513 | 0 | 0 | 1 | 75.35795 | 0 | 0 | 1 | 0.001423 | 0 | 0 | 2 | 0.001858 | 0 | 0 |
| Immune system disorders | 15 | 0.07996923 | 0 | 0 | 1 | 75.35795 | 0 | 0 | 30 | 0.042704 | 0 | 0 | 31 | 0.028805 | 0 | 0 |
| Infections and infestations | 21 | 0.11195692 | 0 | 0 | 0 | 0 | 0 | 0 | 48 | 0.068326 | 0 | 0 | 47 | 0.043673 | 3 | 0.002788 |
| Injury, poisoning and procedural complications | 62 | 0.33053949 | 0 | 0 | 3 | 226.0739 | 0 | 0 | 121 | 0.17224 | 0 | 0 | 113 | 0.105 | 1 | 0.000929 |
| Investigations | 27 | 0.14394462 | 0 | 0 | 0 | 0 | 0 | 0 | 38 | 0.054092 | 0 | 0 | 49 | 0.045531 | 0 | 0 |
| Metabolism and nutrition disorders | 18 | 0.09596308 | 0 | 0 | 0 | 0 | 0 | 0 | 10 | 0.014235 | 0 | 0 | 27 | 0.025088 | 0 | 0 |
| Musculoskeletal and connective tissue disorders | 78 | 0.41584001 | 0 | 0 | 0 | 0 | 0 | 0 | 88 | 0.125265 | 0 | 0 | 36 | 0.033451 | 0 | 0 |
| Neoplasms benign, malignant and unspecified (incl cysts and polyps) | 2 | 0.01066256 | 0 | 0 | 0 | 0 | 0 | 0 | 1 | 0.001423 | 0 | 0 | 5 | 0.004646 | 0 | 0 |
| Nervous system disorders | 322 | 1.71667285 | 0 | 0 | 1 | 75.35795 | 0 | 0 | 303 | 0.431311 | 0 | 0 | 154 | 0.143097 | 0 | 0 |
| Pregnancy, puerperium and perinatal conditions | 4 | 0.02132513 | 0 | 0 | 1 | 75.35795 | 0 | 0 | 0 | 0 | 0 | 0 | 6 | 0.005575 | 0 | 0 |
| Product issues | 18 | 0.09596308 | 0 | 0 | 0 | 0 | 0 | 0 | 118 | 0.167969 | 0 | 0 | 64 | 0.059469 | 0 | 0 |
| Psychiatric disorders | 1542 | 8.22083706 | 2 | 0.010663 | 0 | 0 | 0 | 0 | 174 | 0.247683 | 0 | 0 | 73 | 0.067832 | 1 | 0.000929 |

| | | | | | | | | | | | | | | | | |
|---|---|---|---|---|---|---|---|---|---|---|---|---|---|---|---|---|
| Renal and urinary disorders | 11 | 0.0586441 | 0 | 0 | 0 | 0 | 0 | 0 | 3 | 0.00427 | 0 | 0 | 3 | 0.002788 | 0 | 0 |
| Reproductive system and breast disorders | 2 | 0.01066256 | 0 | 0 | 0 | 0 | 0 | 0 | 7 | 0.009964 | 0 | 0 | 3 | 0.002788 | 0 | 0 |
| Respiratory, thoracic and mediastinal disorders | 128 | 0.68240411 | 0 | 0 | 0 | 0 | 0 | 0 | 587 | 0.835576 | 0 | 0 | 259 | 0.240663 | 1 | 0.000929 |
| Skin and subcutaneous tissue disorders | 149 | 0.79436104 | 0 | 0 | 0 | 0 | 0 | 0 | 177 | 0.251954 | 0 | 0 | 64 | 0.059469 | 0 | 0 |
| Social circumstances | 3 | 0.01599385 | 0 | 0 | 0 | 0 | 0 | 0 | 1 | 0.001423 | 0 | 0 | 5 | 0.004646 | 0 | 0 |
| Surgical and medical procedures | 3 | 0.01599385 | 0 | 0 | 0 | 0 | 0 | 0 | 1 | 0.001423 | 0 | 0 | 5 | 0.004646 | 0 | 0 |
| Vascular disorders | 8 | 0.04265026 | 0 | 0 | 0 | 0 | 0 | 0 | 16 | 0.022775 | 0 | 0 | 11 | 0.010221 | 0 | 0 |

**Table S3.** Raw Higher Level Group Terms (HLGTs) ADRs/fatalities and relative to 100,000 $R_x$ (indicated by (s)).

| | Montelukast | | | | Zafirlukast | | | | Beclometasone | | | | Salbutamol | | | |
|---|---|---|---|---|---|---|---|---|---|---|---|---|---|---|---|---|
| <u>Organ class</u> | All | All (s) | Fatal | Fatal (s) | All | All (s) | Fatal | Fatal (s) | All | All (s) | Fatal | Fatal (s) | All | All (s) | Fatal | Fatal (s) |
| **Blood and lymphatic system disorders** | 9 | 0.047982 | 0 | 0 | 0 | 0 | 0 | 0 | 5 | 0.007117 | 0 | 0 | 2 | 0.001858 | 0 | 0 |
| Anaemias nonhaemolytic and marrow depression | 0 | 0 | 0 | 0 | 0 | 0 | 0 | 0 | 0 | 0 | 0 | 0 | 2 | 0.001858 | | 0 |
| Coagulopathies and bleeding diatheses (excl thrombocytopenic) | 2 | 0.010663 | 0 | 0 | 0 | 0 | 0 | 0 | 1 | 0.001423 | 0 | 0 | 0 | 0 | | 0 |
| Haematological disorders NEC | 2 | 0.010663 | 0 | 0 | 0 | 0 | 0 | 0 | 0 | 0 | 0 | 0 | 0 | 0 | | 0 |
| Haemolyses and related conditions | 1 | 0.005331 | 0 | 0 | 0 | 0 | 0 | 0 | 0 | 0 | 0 | 0 | 0 | 0 | | 0 |
| Platelet disorders | 0 | 0 | 0 | 0 | 0 | 0 | 0 | 0 | 0 | 0 | 0 | 0 | 0 | 0 | | 0 |
| Red blood cell disorders | 1 | 0.005331 | 0 | 0 | 0 | 0 | 0 | 0 | 0 | 0 | 0 | 0 | 0 | 0 | | 0 |
| Spleen, lymphatic and reticuloendothelial system disorders | 2 | 0.010663 | 0 | 0 | 0 | 0 | 0 | 0 | 4 | 0.005694 | 0 | 0 | 0 | 0 | | 0 |
| White blood cell disorders | 1 | 0.005331 | 0 | 0 | 0 | 0 | 0 | 0 | 0 | 0 | 0 | 0 | 0 | 0 | | 0 |
| **Cardiac disorders** | 26 | 0.138613 | 0 | 0 | | 0 | | 0 | | 0 | | 0 | | 0 | | 0 |
| Cardiac arrhythmias | 2 | 0.010663 | 0 | 0 | 0 | 0 | 0 | 0 | 7 | 0.009964 | 0 | 0 | 34 | 0.031593 | | 0 |
| Cardiac disorders, signs and symptoms NEC | 23 | 0.122619 | 0 | 0 | 0 | 0 | 0 | 0 | 60 | 0.085408 | 0 | 0 | 25 | 0.02323 | | 0 |
| Coronary artery disorders | 0 | 0 | 0 | 0 | 0 | 0 | 0 | 0 | 3 | 0.00427 | 0 | 0 | 5 | 0.004646 | | 0 |
| Heart failures | 0 | 0 | 0 | 0 | 0 | 0 | 0 | 0 | 0 | 0 | 0 | 0 | 1 | 0.000929 | | 0 |
| Myocardial disorders | 0 | 0 | 0 | 0 | 0 | 0 | 0 | 0 | 1 | 0.001423 | 0 | 0 | 1 | 0.000929 | | 0 |
| Pericardial disorders | 1 | 0.005331 | 0 | 0 | 0 | 0 | 0 | 0 | 0 | 0 | 0 | 0 | 2 | 0.001858 | | 0 |
| **Congenital, familial and genetic disorders** | 3 | 0.015994 | 0 | 0 | | 0 | | 0 | | 0 | | 0 | | 0 | | 0 |
| Blood and lymphatic system disorders congenital | 0 | 0 | 0 | 0 | 0 | 0 | 0 | 0 | 0 | 0 | 0 | 0 | 0 | 0 | | 0 |
| Cardiac and vascular disorders congenital | 0 | 0 | 0 | 0 | 0 | 0 | 0 | 0 | 0 | 0 | 0 | 0 | 3 | 0.002788 | | 0 |
| Congenital and hereditary disorders NEC | 0 | 0 | 0 | 0 | 0 | 0 | 0 | 0 | 0 | 0 | 0 | 0 | 0 | 0 | | 0 |
| Musculoskeletal and connective tissue disorders congenital | 0 | 0 | 0 | 0 | 0 | 0 | 0 | 0 | 0 | 0 | 0 | 0 | 0 | 0 | | 0 |
| Neurological disorders congenital | 3 | 0.015994 | 0 | 0 | 0 | 0 | 0 | 0 | 0 | 0 | 0 | 0 | 1 | 0.000929 | | 0 |
| Skin and subcutaneous tissue disorders congenital | 0 | 0 | 0 | 0 | 0 | 0 | 0 | 0 | 0 | 0 | 0 | 0 | 0 | 0 | | 0 |
| Reproductive tract and breast disorders congenital | 0 | 0 | 0 | 0 | 0 | 0 | 0 | 0 | 1 | 0.001423 | 0 | 0 | 0 | 0 | | 0 |
| **Ear and labyrinth disorders** | 9 | 0.047982 | 0 | 0 | | 0 | | 0 | | 0 | | 0 | | 0 | | 0 |
| Aural disorders NEC | 3 | 0.015994 | 0 | 0 | 0 | 0 | 0 | 0 | 3 | 0.00427 | 0 | 0 | 1 | 0.000929 | | 0 |
| Hearing disorders | 2 | 0.010663 | 0 | 0 | 0 | 0 | 0 | 0 | 1 | 0.001423 | 0 | 0 | 0 | 0 | | 0 |

| | | | | | | | | | | | | | | | | |
|---|---|---|---|---|---|---|---|---|---|---|---|---|---|---|---|---|
| Inner ear and VIIIth cranial nerve disorders | 4 | 0.021325 | 0 | 0 | 0 | 0 | 0 | 0 | 6 | 0.008541 | 0 | 0 | 4 | 0.003717 | | 0 |
| *Middle ear disorders (excl congenital)* | 0 | 0 | 0 | 0 | 0 | 0 | 0 | 0 | 1 | 0.001423 | 0 | 0 | 0 | 0 | | 0 |
| **Endocrine disorders** | 3 | 0.015994 | | 0 | | 0 | | 0 | | 0 | | 0 | | 0 | | 0 |
| Adrenal gland disorders | 2 | 0.010663 | 0 | 0 | 0 | 0 | | 0 | 2 | 0.002847 | 0 | 0 | 0 | 0 | | 0 |
| Hypothalamus and pituitary gland disorders | 0 | 0 | 0 | 0 | 0 | 0 | | 0 | 1 | 0.001423 | 0 | 0 | 0 | 0 | | 0 |
| Thyroid gland disorders | 1 | 0.005331 | 0 | 0 | 0 | 0 | | 0 | 0 | 0 | 0 | 0 | 1 | 0.000929 | | 0 |
| **Eye disorders** | 22 | 0.117288 | 0 | 0 | | 0 | | 0 | | 0 | | 0 | | 0 | | 0 |
| Anterior eye structural change, deposit and degeneration | 0 | 0 | 0 | 0 | 0 | 0 | 0 | 0 | 5 | 0.007117 | 0 | 0 | 4 | 0.003717 | | 0 |
| Eye disorders NEC | 5 | 0.026656 | 0 | 0 | 0 | 0 | 0 | 0 | 21 | 0.029893 | 0 | 0 | 6 | 0.005575 | | 0 |
| ***Glaucoma and ocular hypertension*** | 0 | 0 | 0 | 0 | 0 | 0 | 0 | 0 | 3 | 0.00427 | 0 | 0 | 3 | 0.002788 | | 0 |
| ***Ocular haemorrhages and vascular disorders NEC*** | 0 | 0 | 0 | 0 | 0 | 0 | 0 | 0 | 2 | 0.002847 | 0 | 0 | 3 | 0.002788 | | 0 |
| Ocular infections, irritations and inflammations | 3 | 0.015994 | 0 | 0 | 0 | 0 | 0 | 0 | 16 | 0.022775 | 0 | 0 | 0 | 0 | | 0 |
| Ocular neuromuscular disorders | 6 | 0.031988 | 0 | 0 | 0 | 0 | 0 | 0 | 1 | 0.001423 | 0 | 0 | 1 | 0.000929 | | 0 |
| Ocular sensory symptoms NEC | 0 | 0 | 0 | 0 | 0 | 0 | 0 | 0 | 5 | 0.007117 | 0 | 0 | 0 | 0 | | 0 |
| Ocular structural change, deposit and degeneration NEC | 1 | 0.005331 | 0 | 0 | 0 | 0 | 0 | 0 | 0 | 0 | 0 | 0 | 0 | 0 | | 0 |
| ***Retina, choroid and vitreous haemorrhages and vascular disorders*** | 0 | 0 | 0 | 0 | 0 | 0 | 0 | 0 | 6 | 0.008541 | 0 | 0 | 0 | 0 | | 0 |
| Vision disorders | 7 | 0.037319 | 0 | 0 | 0 | 0 | 0 | 0 | 26 | 0.03701 | 0 | 0 | 7 | 0.006504 | | 0 |
| **Gastrointestinal disorders** | 204 | 1.087582 | 0 | 0 | | 0 | | 0 | | 0 | | 0 | | 0 | | 0 |
| ***Abdominal hernias and other abdominal wall conditions*** | 0 | 0 | 0 | 0 | 0 | 0 | 0 | 0 | 0 | 0 | 0 | 0 | 1 | 0.000929 | | 0 |
| Anal and rectal conditions NEC | 0 | 0 | 0 | 0 | 0 | 0 | 0 | 0 | 0 | 0 | 0 | 0 | 0 | 0 | | 0 |
| ***Benign neoplasms gastrointestinal*** | 0 | 0 | 0 | 0 | 0 | 0 | 0 | 0 | 1 | 0.001423 | 0 | 0 | 0 | 0 | | 0 |
| Dental and gingival conditions | 2 | 0.010663 | 0 | 0 | 0 | 0 | 0 | 0 | 8 | 0.011388 | 0 | 0 | 2 | 0.001858 | | 0 |
| ***Diverticular disorders*** | 0 | 0 | 0 | 0 | 0 | 0 | 0 | 0 | 0 | 0 | 0 | 0 | 1 | 0.000929 | | 0 |
| Exocrine pancreas conditions | 0 | 0 | 0 | 0 | 0 | 0 | 0 | 0 | 0 | 0 | 0 | 0 | 0 | 0 | | 0 |
| Gastrointestinal conditions NEC | 2 | 0.010663 | 0 | 0 | 0 | 0 | 0 | 0 | 0 | 0 | 0 | 0 | 1 | 0.000929 | | 0 |
| Gastrointestinal haemorrhages NEC | 1 | 0.005331 | 0 | 0 | 0 | 0 | 0 | 0 | 0 | 0 | 0 | 0 | 0 | 0 | | 0 |
| Gastrointestinal inflammatory conditions | 2 | 0.010663 | 0 | 0 | 0 | 0 | 0 | 0 | 2 | 0.002847 | 0 | 0 | 1 | 0.000929 | | 0 |
| Gastrointestinal motility and defaecation conditions | 38 | 0.202589 | 0 | 0 | 0 | 0 | 0 | 0 | 23 | 0.03274 | 0 | 0 | 13 | 0.01208 | | 0 |
| Gastrointestinal signs and symptoms | 122 | 0.650416 | 0 | 0 | 0 | 0 | 0 | 0 | 72 | 0.10249 | 0 | 0 | 28 | 0.026018 | | 0 |
| ***Gastrointestinal stenosis and obstruction*** | 0 | 0 | 0 | 0 | 0 | 0 | 0 | 0 | 1 | 0.001423 | 0 | 0 | 0 | 0 | | 0 |
| Gastrointestinal ulceration and perforation | 0 | 0 | 0 | 0 | 0 | 0 | 0 | 0 | 0 | 0 | 0 | 0 | 0 | 0 | | 0 |
| ***Gastrointestinal vascular conditions*** | 0 | 0 | 0 | 0 | 0 | 0 | 0 | 0 | 0 | 0 | 0 | 0 | 1 | 0.000929 | | 0 |
| Malabsorption conditions | 1 | 0.005331 | 0 | 0 | 0 | 0 | 0 | 0 | 0 | 0 | 0 | 0 | 0 | 0 | | 0 |
| Oral soft tissue conditions | 20 | 0.106626 | 0 | 0 | 0 | 0 | 0 | 0 | 76 | 0.108184 | 0 | 0 | 20 | 0.018584 | | 0 |

| | | | | | | | | | | | | | | | | |
|---|---|---|---|---|---|---|---|---|---|---|---|---|---|---|---|---|
| Peritoneal and retroperitoneal conditions | 0 | 0 | 0 | 0 | 0 | 0 | 0 | 0 | 0 | 0 | 0 | 0 | 0 | 0 | | 0 |
| Salivary gland conditions | 7 | 0.037319 | 0 | 0 | 0 | 0 | 0 | 0 | 11 | 0.015658 | 0 | 0 | 3 | 0.002788 | | 0 |
| Tongue conditions | 9 | 0.047982 | 0 | 0 | 0 | 0 | 0 | 0 | 27 | 0.038434 | 0 | 0 | 4 | 0.003717 | | 0 |
| **General disorders and administration site conditions** | 242 | 1.29017 | 0 | 0 | | 0 | | 0 | | 0 | | 0 | | 0 | | 0 |
| Administration site reactions | 1 | 0.005331 | 0 | 0 | 0 | 0 | 0 | 0 | 7 | 0.009964 | 0 | 0 | 2 | 0.001858 | | 0 |
| Body temperature conditions | 9 | 0.047982 | 0 | 0 | 0 | 0 | 0 | 0 | 9 | 0.012811 | 0 | 0 | 3 | 0.002788 | | 0 |
| *Complications associated with device* | 0 | 0 | 0 | 0 | 0 | 0 | 0 | 0 | 2 | 0.002847 | 0 | 0 | 0 | 0 | | 0 |
| Fatal outcomes | 0 | 0 | 0 | 0 | 0 | 0 | 0 | 0 | 0 | 0 | 0 | 0 | 9 | 0.008363 | | 0 |
| General system disorders NEC | 179 | 0.9543 | 0 | 0 | 0 | 0 | 0 | 0 | 182 | 0.259071 | 1 | 0.001423 | 133 | 0.123584 | | 0 |
| Therapeutic and nontherapeutic effects (excl toxicity) | 53 | 0.282558 | 0 | 0 | 2 | 150.7159 | 0 | 0 | 50 | 0.071173 | 0 | 0 | 59 | 0.054823 | | 0 |
| Tissue disorders NEC | 0 | 0 | 0 | 0 | 0 | 0 | 0 | 0 | 1 | 0.001423 | 0 | 0 | 0 | 0 | | 0 |
| **Hepatobiliary disorders** | 4 | 0.021325 | 0 | 0 | | 0 | | 0 | | 0 | | 0 | | 0 | | 0 |
| Gallbladder disorders | 0 | 0 | 0 | 0 | 0 | 0 | 0 | 0 | 1 | 0.001423 | 0 | 0 | 1 | 0.000929 | | 0 |
| Hepatic and hepatobiliary disorders | 4 | 0.021325 | 0 | 0 | 0 | 0 | 0 | 0 | 0 | 0 | 0 | 0 | 1 | 0.000929 | | 0 |
| **Immune system disorders** | 15 | 0.079969 | 0 | 0 | | 0 | | 0 | | 0 | | 0 | | 0 | | 0 |
| Allergic conditions | 12 | 0.063975 | 0 | 0 | 1 | 75.35795 | 0 | 0 | 29 | 0.041281 | 0 | 0 | 28 | 0.026018 | | 0 |
| *Autoimmune disorders* | 0 | 0 | 0 | 0 | 0 | 0 | 0 | 0 | 1 | 0.001423 | 0 | 0 | 2 | 0.001858 | | 0 |
| Immune disorders NEC | 3 | 0.015994 | 0 | 0 | 0 | 0 | 0 | 0 | 0 | 0 | 0 | 0 | 1 | 0.000929 | | 0 |
| **Infections and infestations** | 21 | 0.111957 | 0 | 0 | | 0 | | 0 | | 0 | | 0 | | 0 | | 0 |
| Ancillary infectious topics | 0 | 0 | 0 | 0 | 0 | 0 | 0 | 0 | 0 | 0 | 0 | 0 | 0 | 0 | | 0 |
| Bacterial infectious disorders | 0 | 0 | 0 | 0 | 0 | 0 | 0 | 0 | 1 | 0.001423 | 0 | 0 | 1 | 0.000929 | | 0 |
| Fungal infectious disorders | 4 | 0.021325 | 0 | 0 | 0 | 0 | 0 | 0 | 21 | 0.029893 | 0 | 0 | 9 | 0.008363 | | 0 |
| Infections - pathogen unspecified | 13 | 0.069307 | 0 | 0 | 1 | 75.35795 | 0 | 0 | 15 | 0.021352 | 0 | 0 | 30 | 0.027876 | | 0 |
| Mycoplasmal infectious disorders | 0 | 0 | 0 | 0 | 0 | 0 | 0 | 0 | 0 | 0 | 0 | 0 | 0 | 0 | | 0 |
| Viral infectious disorders | 4 | 0.021325 | 0 | 0 | 0 | 0 | 0 | 0 | 11 | 0.015658 | 0 | 0 | 7 | 0.006504 | | 0 |
| **Injury, poisoning and procedural complications** | 62 | 0.330539 | 0 | 0 | | 0 | | 0 | | 0 | | 0 | | 0 | | 0 |
| Bone and joint injuries | 0 | 0 | 0 | 0 | 0 | 0 | 0 | 0 | 2 | 0.002847 | 0 | 0 | 0 | 0 | | 0 |
| Exposures, chemical injuries and poisoning | 2 | 0.010663 | 0 | 0 | 1 | 75.35795 | 0 | 0 | 5 | 0.007117 | 0 | 0 | 17 | 0.015796 | | 0 |
| Injuries NEC | 14 | 0.074638 | 0 | 0 | 0 | 0 | 0 | 0 | 19 | 0.027046 | 0 | 0 | 11 | 0.010221 | | 0 |
| Medication errors and other product use errors and issues | 41 | 0.218583 | 0 | 0 | 1 | 75.35795 | 0 | 0 | 82 | 0.116724 | 0 | 0 | 64 | 0.059469 | | 0 |
| Off label uses and intentional product misuses/use issues | 3 | 0.015994 | 0 | 0 | 1 | 75.35795 | 0 | 0 | 8 | 0.011388 | 0 | 0 | 10 | 0.009292 | | 0 |
| Overdoses and underdoses NEC | 2 | 0.010663 | 0 | 0 | 0 | 0 | 0 | 0 | 4 | 0.005694 | 0 | 0 | 11 | 0.010221 | | 0 |

| | | | | | | | | | | | | | | | | |
|---|---|---|---|---|---|---|---|---|---|---|---|---|---|---|---|---|
| Procedural related injuries and complications NEC | 0 | 0 | 0 | 0 | 0 | 0 | 0 | 0 | 1 | 0.001423 | 0 | 0 | 0 | 0 | | 0 |
| **Investigations** | 27 | 0.143945 | 0 | 0 | | 0 | | 0 | | 0 | | 0 | | 0 | | 0 |
| Cardiac and vascular investigations (excl enzyme tests) | 7 | 0.037319 | 0 | 0 | 0 | 0 | 0 | 0 | 17 | 0.024199 | 0 | 0 | 10 | 0.009292 | | 0 |
| ***Cytogenetic investigations and genetic analyses*** | 0 | 0 | 0 | 0 | 0 | 0 | 0 | 0 | 1 | 0.001423 | 0 | 0 | 0 | 0 | | 0 |
| Enzyme investigations NEC | 1 | 0.005331 | 0 | 0 | 0 | 0 | 0 | 0 | 0 | 0 | 0 | 0 | 0 | 0 | | 0 |
| ***Gastrointestinal investigations*** | 0 | 0 | 0 | 0 | 0 | 0 | 0 | 0 | 0 | 0 | 0 | 0 | 1 | 0.000929 | | 0 |
| Haematology investigations (incl blood groups) | 0 | 0 | 0 | 0 | 0 | 0 | 0 | 0 | 0 | 0 | 0 | 0 | 2 | 0.001858 | | 0 |
| Hepatobiliary investigations | 3 | 0.015994 | 0 | 0 | 0 | 0 | 0 | 0 | 0 | 0 | 0 | 0 | 2 | 0.001858 | | 0 |
| Lipid analyses | 0 | 0 | 0 | 0 | 0 | 0 | 0 | 0 | 0 | 0 | 0 | 0 | 2 | 0.001858 | | 0 |
| Immunology and allergy investigations | 0 | 0 | 0 | 0 | 0 | 0 | 0 | 0 | 0 | 0 | 0 | 0 | 0 | 0 | | 0 |
| Investigations, imaging and histopathology procedures NEC | 1 | 0.005331 | 0 | 0 | 0 | 0 | 0 | 0 | 0 | 0 | 0 | 0 | 0 | 0 | | 0 |
| Metabolic, nutritional and blood gas investigations | 6 | 0.031988 | 0 | 0 | 0 | 0 | 0 | 0 | 3 | 0.00427 | 0 | 0 | 18 | 0.016726 | | 0 |
| Microbiology and serology investigations | 0 | 0 | 0 | 0 | 0 | 0 | 0 | 0 | 0 | 0 | 0 | 0 | 1 | 0.000929 | | 0 |
| ***Neurological, special senses and psychiatric investigations*** | 0 | 0 | 0 | 0 | 0 | 0 | 0 | 0 | 2 | 0.002847 | 0 | 0 | 2 | 0.001858 | | 0 |
| ***Protein and chemistry analyses NEC*** | 0 | 0 | 0 | 0 | 0 | 0 | 0 | 0 | 2 | 0.002847 | 0 | 0 | 0 | 0 | | 0 |
| Physical examination and organ system status topics | 7 | 0.037319 | 0 | 0 | 0 | 0 | 0 | 0 | 8 | 0.011388 | 0 | 0 | 5 | 0.004646 | | 0 |
| Renal and urinary tract investigations and urinalyses | 0 | 0 | 0 | 0 | 0 | 0 | 0 | 0 | 1 | 0.001423 | 0 | 0 | 0 | 0 | | 0 |
| Respiratory and pulmonary investigations (excl blood gases) | 1 | 0.005331 | 0 | 0 | 0 | 0 | 0 | 0 | 3 | 0.00427 | 0 | 0 | 7 | 0.006504 | | 0 |
| Toxicology and therapeutic drug monitoring | 1 | 0.005331 | 0 | 0 | 0 | 0 | 0 | 0 | 1 | 0.001423 | 0 | 0 | 0 | 0 | | 0 |
| **Metabolism and nutrition disorders** | 18 | 0.095963 | 0 | 0 | 0 | 0 | | 0 | | 0 | | 0 | | 0 | | 0 |
| Acid-base disorders | 0 | 0 | 0 | 0 | 0 | 0 | 0 | 0 | 0 | 0 | 0 | 0 | 9 | 0.008363 | | 0 |
| Appetite and general nutritional disorders | 14 | 0.074638 | 0 | 0 | 0 | 0 | 0 | 0 | 6 | 0.008541 | 0 | 0 | 4 | 0.003717 | | 0 |
| Bone, calcium, magnesium and phosphorus metabolism disorders | 0 | 0 | 0 | 0 | 0 | 0 | 0 | 0 | 0 | 0 | 0 | 0 | 1 | 0.000929 | | 0 |
| Electrolyte and fluid balance conditions | 3 | 0.015994 | 0 | 0 | 0 | 0 | 0 | 0 | 1 | 0.001423 | 0 | 0 | 11 | 0.010221 | | 0 |
| Food intolerance syndromes | 0 | 0 | 0 | 0 | 0 | 0 | 0 | 0 | 0 | 0 | 0 | 0 | | 0 | | 0 |
| Glucose metabolism disorders (incl diabetes mellitus) | 0 | 0 | 0 | 0 | 0 | 0 | 0 | 0 | 3 | 0.00427 | 0 | 0 | 1 | 0.000929 | | 0 |
| Lipid metabolism disorders | 1 | 0.005331 | 0 | 0 | 0 | 0 | 0 | 0 | 0 | 0 | 0 | 0 | | 0 | | 0 |
| Purine and pyrimidine metabolism disorders | 0 | 0 | 0 | 0 | 0 | 0 | 0 | 0 | 0 | 0 | 0 | 0 | | 0 | | 0 |

| | | | | | | | | | | | | | | | | |
|---|---|---|---|---|---|---|---|---|---|---|---|---|---|---|---|---|
| ***Vitamin related disorders*** | 0 | 0 | 0 | 0 | 0 | 0 | 0 | 0 | 0 | 0 | 0 | 0 | 1 | 0.000929 | | 0 |
| **Musculoskeletal and connective tissue disorders** | 78 | 0.41584 | 0 | 0 | | 0 | | 0 | | 0 | | 0 | | 0 | | 0 |
| Bone disorders (excl congenital and fractures) | 1 | 0.005331 | 0 | 0 | 0 | 0 | 0 | 0 | 5 | 0.007117 | 0 | 0 | 0 | 0 | | 0 |
| Connective tissue disorders (excl congenital) | 1 | 0.005331 | 0 | 0 | 0 | 0 | 0 | 0 | 1 | 0.001423 | 0 | 0 | 0 | 0 | | 0 |
| Fractures | 0 | 0 | 0 | 0 | 0 | 0 | 0 | 0 | 0 | 0 | 0 | 0 | 0 | 0 | | 0 |
| Joint disorders | 25 | 0.133282 | 0 | 0 | 0 | 0 | 0 | 0 | 13 | 0.018505 | 0 | 0 | 10 | 0.009292 | | 0 |
| Muscle disorders | 39 | 0.20792 | 0 | 0 | 0 | 0 | 0 | 0 | 53 | 0.075444 | 0 | 0 | 17 | 0.015796 | | 0 |
| ***Musculoskeletal and connective tissue deformities (incl intervertebral disc disorders)*** | 0 | 0 | 0 | 0 | 0 | 0 | 0 | 0 | 0 | 0 | 0 | 0 | 1 | 0.000929 | | 0 |
| Musculoskeletal and connective tissue disorders NEC | 12 | 0.063975 | 0 | 0 | 0 | 0 | 0 | 0 | 14 | 0.019929 | 0 | 0 | 8 | 0.007434 | | 0 |
| Synovial and bursal disorders | 0 | 0 | 0 | 0 | 0 | 0 | 0 | 0 | 0 | 0 | 0 | 0 | 0 | 0 | | 0 |
| Tendon, ligament and cartilage disorders | 0 | 0 | 0 | 0 | 0 | 0 | 0 | 0 | 2 | 0.002847 | 0 | 0 | 0 | 0 | | 0 |
| **Neoplasms benign, malignant and unspecified (incl cysts and polyps)** | 2 | 0.010663 | 0 | 0 | | 0 | | 0 | | 0 | | 0 | | 0 | | 0 |
| Breast neoplasms benign (incl nipple) | 1 | 0.005331 | 0 | 0 | 0 | 0 | 0 | 0 | 0 | 0 | 0 | 0 | 0 | 0 | | 0 |
| Cutaneous neoplasms benign | 1 | 0.005331 | 0 | 0 | 0 | 0 | 0 | 0 | 0 | 0 | 0 | 0 | 1 | 0.000929 | | 0 |
| Gastrointestinal neoplasms malignant and unspecified | 0 | 0 | 0 | 0 | 0 | 0 | 0 | 0 | 0 | 0 | 0 | 0 | 0 | 0 | | 0 |
| ***Miscellaneous and site unspecified neoplasms benign*** | 0 | 0 | 0 | 0 | 0 | 0 | 0 | 0 | 0 | 0 | 0 | 0 | 1 | 0.000929 | | 0 |
| ***Miscellaneous and site unspecified neoplasms malignant and unspecified*** | 0 | 0 | 0 | 0 | 0 | 0 | 0 | 0 | 0 | 0 | 0 | 0 | 2 | 0.001858 | | 0 |
| ***Respiratory and mediastinal neoplasms malignant and unspecified*** | 0 | 0 | 0 | 0 | 0 | 0 | 0 | 0 | 0 | 0 | 0 | 0 | 1 | 0.000929 | | 0 |
| Leukaemias | 0 | 0 | 0 | 0 | 0 | 0 | 0 | 0 | 0 | 0 | 0 | 0 | 0 | 0 | | 0 |
| ***Plasma cell neoplasms*** | | 0 | | 0 | | 0 | | 0 | 1 | 0.001423 | 0 | 0 | 0 | 0 | | 0 |
| Skin neoplasms malignant and unspecified | 0 | 0 | 0 | 0 | 0 | 0 | 0 | 0 | 0 | 0 | 0 | 0 | 0 | 0 | | 0 |
| **Nervous system disorders** | 322 | 1.716673 | 0 | 0 | | 0 | | 0 | | 0 | | 0 | | 0 | | 0 |
| Central nervous system vascular disorders | 0 | 0 | 0 | 0 | 0 | 0 | 0 | 0 | 0 | 0 | 0 | 0 | 2 | 0.001858 | | 0 |
| Cranial nerve disorders (excl neoplasms) | 1 | 0.005331 | 0 | 0 | 0 | 0 | 0 | 0 | 6 | 0.008541 | 0 | 0 | 2 | 0.001858 | | 0 |
| Demyelinating disorders | 0 | 0 | 0 | 0 | 0 | 0 | 0 | 0 | 0 | 0 | 0 | 0 | 1 | 0.000929 | | 0 |
| Encephalopathies | 0 | 0 | 0 | 0 | 0 | 0 | 0 | 0 | 0 | 0 | 0 | 0 | 0 | 0 | | 0 |
| Headaches | 85 | 0.453159 | 0 | 0 | 0 | 0 | 0 | 0 | 91 | 0.129536 | 0 | 0 | 23 | 0.021372 | | 0 |
| Increased intracranial pressure and hydrocephalus | 0 | 0 | 0 | 0 | 0 | 0 | 0 | 0 | 0 | 0 | 0 | 0 | 0 | 0 | | 0 |
| Mental impairment disorders | 46 | 0.245239 | 0 | 0 | 0 | 0 | 0 | 0 | 5 | 0.007117 | 0 | 0 | 5 | 0.004646 | | 0 |

| | | | | | | | | | | | | | | | | |
|---|---|---|---|---|---|---|---|---|---|---|---|---|---|---|---|---|
| Movement disorders (incl parkinsonism) | 53 | 0.282558 | 0 | 0 | 0 | 0 | 0 | 0 | 87 | 0.123842 | 0 | 0 | 47 | 0.043673 | | 0 |
| Nervous system neoplasms benign | 0 | 0 | 0 | 0 | 0 | 0 | 0 | 0 | 0 | 0 | 0 | 0 | 1 | 0.000929 | | 0 |
| Neurological disorders NEC | 121 | 0.645085 | 0 | 0 | 1 | 75.35795 | 0 | 0 | 110 | 0.156581 | 0 | 0 | 59 | 0.054823 | | 0 |
| Neurological disorders of the eye | 0 | 0 | 0 | 0 | 0 | 0 | 0 | 0 | 0 | 0 | 0 | 0 | 0 | 0 | | 0 |
| Neuromuscular disorders | 0 | 0 | 0 | 0 | 0 | 0 | 0 | 0 | 0 | 0 | 0 | 0 | 7 | 0.006504 | | 0 |
| Peripheral neuropathies | 4 | 0.021325 | 0 | 0 | 0 | 0 | 0 | 0 | 1 | 0.001423 | 0 | 0 | 3 | 0.002788 | | 0 |
| Seizures (incl subtypes) | 10 | 0.053313 | 0 | 0 | 0 | 0 | 0 | 0 | 2 | 0.002847 | 0 | 0 | 0 | 0 | | 0 |
| Sleep disturbances (incl subtypes) | 2 | 0.010663 | 0 | 0 | 0 | 0 | 0 | 0 | 0 | 0 | 0 | 0 | 2 | 0.001858 | | 0 |
| Spinal cord and nerve root disorders | 0 | 0 | 0 | 0 | 0 | 0 | 0 | 0 | 1 | 0.001423 | 0 | 0 | 1 | 0.000929 | | 0 |
| Structural brain disorders | 0 | 0 | 0 | 0 | 0 | 0 | 0 | 0 | 0 | 0 | 0 | 0 | 1 | 0.000929 | | 0 |
| **Pregnancy, puerperium and perinatal conditions** | 4 | 0.021325 | 0 | 0 | | 0 | | 0 | | 0 | | 0 | 6 | 0.005575 | | 0 |
| Abortions and stillbirth | 0 | 0 | 0 | 0 | 1 | 75.35795 | 0 | 0 | 0 | 0 | 0 | 0 | 0 | 0 | | 0 |
| Foetal complications | 1 | 0.005331 | 0 | 0 | 0 | 0 | 0 | 0 | 0 | 0 | 0 | 0 | 0 | 0 | | 0 |
| Maternal complications of labour and delivery | 2 | 0.010663 | 0 | 0 | 0 | 0 | 0 | 0 | 0 | 0 | 0 | 0 | 0 | 0 | | 0 |
| Maternal complications of pregnancy | 0 | 0 | 0 | 0 | 0 | 0 | 0 | 0 | 0 | 0 | 0 | 0 | 2 | 0.001858 | | 0 |
| Neonatal and perinatal conditions | 1 | 0.005331 | 0 | 0 | 0 | 0 | 0 | 0 | 0 | 0 | 0 | 0 | 3 | 0.002788 | | 0 |
| Pregnancy, labour, delivery and postpartum conditions | 0 | 0 | 0 | 0 | 0 | 0 | 0 | 0 | 0 | 0 | 0 | 0 | 1 | 0.000929 | | 0 |
| **Product issues** | 18 | 0.095963 | 0 | 0 | | 0 | | 0 | | 0 | | 0 | 64 | 0.059469 | | 0 |
| Device issues | 0 | 0 | 0 | 0 | 0 | 0 | 0 | 0 | 51 | 0.072597 | 0 | 0 | 10 | 0.009292 | | 0 |
| Product quality, supply, distribution, manufacturing and quality system issues | 18 | 0.095963 | 0 | 0 | 0 | 0 | 0 | 0 | 67 | 0.095372 | 0 | 0 | 54 | 0.050177 | | 0 |
| **Psychiatric disorders** | 1542 | 8.220837 | 2 | 0.010663 | | 0 | | 0 | | 0 | | 0 | 73 | 0.067832 | | 0 |
| Adjustment disorders (incl subtypes) | 0 | 0 | 0 | 0 | 0 | 0 | 0 | 0 | 0 | 0 | 0 | 0 | 0 | 0 | | 0 |
| Anxiety disorders and symptoms | 275 | 1.466103 | 0 | 0 | 0 | 0 | 0 | 0 | 58 | 0.082561 | 0 | 0 | 22 | 0.020442 | | 0 |
| Changes in physical activity | 37 | 0.197257 | 0 | 0 | 0 | 0 | 0 | 0 | 8 | 0.011388 | 0 | 0 | 1 | 0.000929 | | 0 |
| Cognitive and attention disorders and disturbances | 6 | 0.031988 | 0 | 0 | 0 | 0 | 0 | 0 | 1 | 0.001423 | 0 | 0 | 0 | 0 | | 0 |
| Communication disorders and disturbances | 15 | 0.079969 | 0 | 0 | 0 | 0 | 0 | 0 | 1 | 0.001423 | 0 | 0 | 0 | 0 | | 0 |
| Deliria (incl confusion) | 19 | 0.101294 | 0 | 0 | 0 | 0 | 0 | 0 | 5 | 0.007117 | 0 | 0 | 3 | 0.002788 | | 0 |
| Depressed mood disorders and disturbances | 162 | 0.863668 | 0 | 0 | 0 | 0 | 0 | 0 | 17 | 0.024199 | 0 | 0 | 8 | 0.007434 | | 0 |
| Developmental disorders NEC | 1 | 0.005331 | 0 | 0 | 0 | 0 | 0 | 0 | 0 | 0 | 0 | 0 | 0 | 0 | | 0 |
| Dissociative disorders | 5 | 0.026656 | 0 | 0 | 0 | 0 | 0 | 0 | 0 | 0 | 0 | 0 | 0 | 0 | | 0 |
| Disturbances in thinking and perception | 76 | 0.405177 | 0 | 0 | 0 | 0 | 0 | 0 | 5 | 0.007117 | 0 | 0 | 6 | 0.005575 | | 0 |
| Eating disorders and disturbances | 0 | 0 | 0 | 0 | 0 | 0 | 0 | 0 | 0 | 0 | 0 | 0 | 0 | 0 | | 0 |

| | | | | | | | | | | | | | | | | |
|---|---|---|---|---|---|---|---|---|---|---|---|---|---|---|---|---|
| Impulse control disorders NEC | 4 | 0.021325 | 0 | 0 | 0 | 0 | 0 | 0 | 0 | 0 | 0 | 0 | 0 | 0 | | 0 |
| Manic and bipolar mood disorders and disturbances | 6 | 0.031988 | 0 | 0 | 0 | 0 | 0 | 0 | 0 | 0 | 0 | 0 | 0 | 0 | | 0 |
| Mood disorders and disturbances NEC | 182 | 0.970293 | 0 | 0 | 0 | 0 | 0 | 0 | 25 | 0.035587 | 0 | 0 | 5 | 0.004646 | | 0 |
| Personality disorders and disturbances in behaviour | 144 | 0.767705 | 0 | 0 | 0 | 0 | 0 | 0 | 14 | 0.019929 | 0 | 0 | 1 | 0.000929 | | 0 |
| Psychiatric and behavioural symptoms NEC | 65 | 0.346533 | 0 | 0 | 0 | 0 | 0 | 0 | 7 | 0.009964 | 0 | 0 | 1 | 0.000929 | | 0 |
| Psychiatric disorders NEC | 26 | 0.138613 | 0 | 0 | 0 | 0 | 0 | 0 | 0 | 0 | 0 | 0 | 6 | 0.005575 | | 0 |
| Schizophrenia and other psychotic disorders | 12 | 0.063975 | 0 | 0 | 0 | 0 | 0 | 0 | 0 | 0 | 0 | 0 | 1 | 0.000929 | | 0 |
| Sexual dysfunctions, disturbances and gender identity disorders | 0 | 0 | 0 | 0 | 0 | 0 | 0 | 0 | 0 | 0 | 0 | 0 | 1 | 0.000929 | | 0 |
| Sleep disorders and disturbances | 411 | 2.191157 | 0 | 0 | 0 | 0 | 0 | 0 | 30 | 0.042704 | 0 | 0 | 15 | 0.013938 | | 0 |
| ***Somatic symptom and related disorders*** | | | | | | | | | | | | | 1 | 0.000929 | | |
| Suicidal and self-injurious behaviours NEC | 96 | 0.511803 | 2 | 0.010663 | 0 | 0 | 0 | 0 | 3 | 0.00427 | 0 | 0 | 2 | 0.001858 | | 0 |
| **Renal and urinary disorders** | 11 | 0.058644 | | 0 | | 0 | | 0 | | 0 | | 0 | 3 | 0.002788 | | 0 |
| Nephropathies | 0 | 0 | 0 | 0 | 0 | 0 | 0 | 0 | 0 | 0 | 0 | 0 | 0 | 0 | | 0 |
| ***Bladder and bladder neck disorders (excl calculi)*** | | | | | | | | | | | | | 1 | 0.000929 | | |
| Renal disorders (excl nephropathies) | 0 | 0 | 0 | 0 | 0 | 0 | 0 | 0 | 0 | 0 | 0 | 0 | 2 | 0.001858 | | 0 |
| Urethral disorders (excl calculi) | 0 | 0 | 0 | 0 | 0 | 0 | 0 | 0 | 0 | 0 | 0 | 0 | 0 | 0 | | 0 |
| Urinary tract signs and symptoms | 11 | 0.058644 | 0 | 0 | 0 | 0 | 0 | 0 | 3 | 0.00427 | 0 | 0 | 0 | 0 | | 0 |
| **Reproductive system and breast disorders** | 2 | 0.010663 | | 0 | | 0 | | 0 | | 0 | | 0 | | 0 | | 0 |
| Breast disorders | 0 | 0 | 0 | 0 | 0 | 0 | 0 | 0 | 0 | 0 | 0 | 0 | 2 | 0.001858 | | 0 |
| Menstrual cycle and uterine bleeding disorders | 0 | 0 | 0 | 0 | 0 | 0 | 0 | 0 | 5 | 0.007117 | 0 | 0 | 0 | 0 | | 0 |
| Reproductive tract disorders NEC | 0 | 0 | 0 | 0 | 0 | 0 | 0 | 0 | 1 | 0.001423 | 0 | 0 | 0 | 0 | | 0 |
| Sexual function and fertility disorders | 0 | 0 | 0 | 0 | 0 | 0 | 0 | 0 | 0 | 0 | 0 | 0 | 0 | 0 | | 0 |
| Testicular and epididymal disorders | 1 | 0.005331 | 0 | 0 | 0 | 0 | 0 | 0 | 0 | 0 | 0 | 0 | 0 | 0 | | 0 |
| Uterine, pelvic and broad ligament disorders | 1 | 0.005331 | 0 | 0 | 0 | 0 | 0 | 0 | 0 | 0 | 0 | 0 | 0 | 0 | | 0 |
| Vulvovaginal disorders (excl infections and inflammations) | 0 | 0 | 0 | 0 | 0 | 0 | 0 | 0 | 1 | 0.001423 | 0 | 0 | 1 | 0.000929 | | 0 |
| **Respiratory, thoracic and mediastinal disorders** | 128 | | | 0 | | 0 | | 0 | | 0 | | 0 | 259 | 0.240663 | | 0 |
| Bronchial disorders (excl neoplasms) | 21 | 0.111957 | 0 | 0 | 0 | 0 | 0 | 0 | 108 | 0.153735 | 0 | 0 | 75 | 0.06969 | | 0 |
| Lower respiratory tract disorders (excl obstruction and infection) | 3 | 0.015994 | 0 | 0 | 0 | 0 | 0 | 0 | 1 | 0.001423 | 0 | 0 | 3 | 0.002788 | | 0 |
| Pleural disorders | 0 | 0 | 0 | 0 | 0 | 0 | 0 | 0 | 0 | 0 | 0 | 0 | 1 | 0.000929 | | 0 |
| Pulmonary vascular disorders | 0 | 0 | 0 | 0 | 0 | 0 | 0 | 0 | 0 | 0 | 0 | 0 | 0 | 0 | | 0 |

| | | | | | | | | | | | | | | | | |
|---|---|---|---|---|---|---|---|---|---|---|---|---|---|---|---|---|
| Respiratory disorders NEC | 44 | 0.234576 | 0 | 0 | 0 | 0 | 0 | 0 | 265 | 0.377219 | 0 | 0 | 144 | 0.133805 | | 0 |
| Respiratory tract signs and symptoms | 32 | 0.170601 | 0 | 0 | 0 | 0 | 0 | 0 | 142 | 0.202132 | 0 | 0 | 31 | 0.028805 | | 0 |
| Upper respiratory tract disorders (excl infections) | 28 | 0.149276 | 0 | 0 | 0 | 0 | 0 | 0 | 71 | 0.101066 | 0 | 0 | 5 | 0.004646 | | 0 |
| **Skin and subcutaneous tissue disorders** | 149 | 0.794361 | | 0 | | 0 | | 0 | | 0 | | 0 | 64 | 0.059469 | | 0 |
| Angioedema and urticaria | 30 | 0.159938 | 0 | 0 | 0 | 0 | 0 | 0 | 19 | 0.027046 | 0 | 0 | 10 | 0.009292 | | 0 |
| Cornification and dystrophic skin disorders | 0 | 0 | 0 | 0 | 0 | 0 | 0 | 0 | 2 | 0.002847 | 0 | 0 | 0 | 0 | | 0 |
| Epidermal and dermal conditions | 87 | 0.463822 | 0 | 0 | 0 | 0 | 0 | 0 | 141 | 0.200709 | 0 | 0 | 44 | 0.040885 | | 0 |
| Pigmentation disorders | 0 | 0 | 0 | 0 | 0 | 0 | 0 | 0 | 1 | 0.001423 | 0 | 0 | 0 | 0 | | 0 |
| Skin and subcutaneous tissue disorders NEC | 2 | 0.010663 | 0 | 0 | 0 | 0 | 0 | 0 | 0 | 0 | 0 | 0 | 1 | 0.000929 | | 0 |
| Skin appendage conditions | 29 | 0.154607 | 0 | 0 | 0 | 0 | 0 | 0 | 13 | 0.018505 | 0 | 0 | 8 | 0.007434 | | 0 |
| Skin vascular abnormalities | 1 | 0.005331 | 0 | 0 | 0 | 0 | 0 | 0 | 1 | 0.001423 | 0 | 0 | 1 | 0.000929 | | 0 |
| **Social circumstances** | 3 | 0.015994 | | 0 | | 0 | | 0 | | 0 | | 0 | 5 | 0.004646 | | 0 |
| Legal issues | 1 | 0.005331 | 0 | 0 | 0 | 0 | 0 | 0 | 0 | 0 | 0 | 0 | 0 | 0 | | 0 |
| Lifestyle issues | 2 | 0.010663 | 0 | 0 | 0 | 0 | 0 | 0 | 1 | 0.001423 | 0 | 0 | 5 | 0.004646 | | 0 |
| **Surgical and medical procedures** | 3 | 0.015994 | | 0 | | 0 | | 0 | | 0 | | 0 | 5 | 0.004646 | | 0 |
| Bone and joint therapeutic procedures | 0 | 0 | 0 | 0 | 0 | 0 | 0 | 0 | 0 | 0 | 0 | 0 | | 0 | | 0 |
| Gastrointestinal therapeutic procedures | 1 | 0.005331 | 0 | 0 | 0 | 0 | 0 | 0 | 0 | 0 | 0 | 0 | 1 | 0.000929 | | 0 |
| Obstetric and gynaecological therapeutic procedures | 1 | 0.005331 | 0 | 0 | 0 | 0 | 0 | 0 | 0 | 0 | 0 | 0 | 1 | 0.000929 | | 0 |
| Therapeutic procedures and supportive care NEC | 1 | 0.005331 | 0 | 0 | 0 | 0 | 0 | 0 | 1 | 0.001423 | 0 | 0 | 3 | 0.002788 | | 0 |
| **Vascular disorders** | 8 | 0.04265 | | 0 | | 0 | | 0 | | 0 | | 0 | 11 | 0.010221 | | 0 |
| Arteriosclerosis, stenosis, vascular insufficiency and necrosis | 1 | 0.005331 | 0 | 0 | 0 | 0 | 0 | 0 | 1 | 0.001423 | 0 | 0 | 0 | 0 | | 0 |
| Decreased and nonspecific blood pressure disorders and shock | 1 | 0.005331 | 0 | 0 | 0 | 0 | 0 | 0 | 4 | 0.005694 | 0 | 0 | 1 | 0.000929 | | 0 |
| Embolism and thrombosis | 0 | 0 | 0 | 0 | 0 | 0 | 0 | 0 | 4 | 0.005694 | 0 | 0 | 0 | 0 | | 0 |
| Vascular disorders NEC | 3 | 0.015994 | 0 | 0 | 0 | 0 | 0 | 0 | 3 | 0.00427 | 0 | 0 | 4 | 0.003717 | | 0 |
| Vascular haemorrhagic disorders | 0 | 0 | 0 | 0 | 0 | 0 | 0 | 0 | 2 | 0.002847 | 0 | 0 | 0 | 0 | | 0 |
| Vascular hypertensive disorders | 2 | 0.010663 | 0 | 0 | 0 | 0 | 0 | 0 | 1 | 0.001423 | 0 | 0 | 6 | 0.005575 | | 0 |
| Vascular infections and inflammations | 1 | 0.005331 | 0 | 0 | 0 | 0 | 0 | 0 | 0 | 0 | 0 | 0 | 0 | 0 | | 0 |
| Venous varices | 0 | 0 | 0 | 0 | 0 | 0 | 0 | 0 | 1 | 0.001423 | 0 | 0 | 0 | 0 | | 0 |
